# Machine learning framework for predicting the presence of high-risk clonal haematopoiesis using complete blood count data: a population-based study of 431,531 UK Biobank participants

**DOI:** 10.1101/2024.09.30.24314606

**Authors:** William G. Dunn, Isabella Withnell, Muxin Gu, Pedro Quiros, Sruthi Cheloor Kovilakam, Ludovica Marando, Sean Wen, Margarete A Fabre, Irina Mohorianu, Dragana Vuckovic, George S. Vassiliou

**Affiliations:** Cambridge Stem Cell Institute, University of Cambridge, Cambridge, UK; Department of Haematology, University of Cambridge, Cambridge, UK; Department of Haematology, Cambridge University Hospitals NHS Trust, Cambridge, UK; Division of Biosciences, University College London, London, UK; Department of Biochemistry and Molecular Biology, Instituto Universitario de Oncología (IUOPA), Universidad de Oviedo, 33006, Oviedo, Spain; Department of Hematology, Mayo Clinic, Rochester, MI, USA; Centre for Genomics Research, Discovery Sciences, Biopharmaceuticals R&D, AstraZeneca, Cambridge, UK; Department of Epidemiology and Biostatistics, School of Public Health, Faculty of Medicine, Imperial College London, London, UK

## Abstract

**Background:** Clonal haematopoiesis (CH), the disproportionate expansion of a haematopoietic stem cell and its progeny, driven by somatic DNA mutations, is a common age-related phenomenon that engenders an increased risk of developing myeloid neoplasms (MN). At present, CH is identified by targeted sequencing of peripheral blood DNA, which is impractical to apply at population scale. The complete blood count (CBC) is an inexpensive, widely used clinical test. Here, we explore whether machine learning (ML) approaches applied to CBC data could predict individuals likely to harbour CH and prioritise them for DNA sequencing.

**Methods:** The UK Biobank was filtered to identify 431,531 participants with paired CBC and whole exome sequencing (WES). Somatic mutations were previously identified from blood WES using Mutect2 to classify individuals with CH driver mutations. Using 18 CBC indices/features and basic demographics (age and sex), we trained a range of tree-based ML classifiers to infer as binary output, the presence/ absence of CH.

**Findings:** Using Random Forest (RF) classifiers, we predicted the presence/absence of CH driven by mutations in one of five genes known to confer a high-risk of incident MN (*JAK2, CALR, SF3B1, SRSF2* and *U2AF1*). We subsequently developed a unified, optimised RF classifier for high-risk CH driven by any of these genes and assessed its performance (median AUC 0.85). However, the low prevalence of high-risk CH implies that our model cannot be generalised to population scale without compromising its sensitivity (20.1% using stringent cutoff probability score).

**Interpretation:** We showcase a proof-of-concept that the presence of high-risk CH can be inferred from CBC perturbations using RF classifiers. The future integration of raw blood cell analyser data can help improve the performance of our model and facilitate its application at scale.

**Funding:** Cancer Research UK.

**Research in context:** 

**Evidence before this study:** We searched PubMed for articles published, in English, between database inception and 5^th^ of June 2024, using the terms “clonal hematopoiesis” AND (“machine learning” OR “artificial intelligence”). We additionally searched for the terms “clonal hematopoesis” AND “complete blood count”. We found 18 research articles: one article used ML approaches (XGBoost classifiers) to differentiate clonal haematopoiesis “driver” mutations from “passenger” mutations, but none linked machine learning frameworks to complete blood count data for predicting the presence of clonal haematopoiesis. Progression from clonal haematopoiesis to myeloid neoplasia is known to be associated with several blood count parameters; two recent publications developed clonal haematopoiesis risk stratification tools that incorporated blood count indices in their final risk prediction models (Gu et al. Nature Genetics, Weeks et al. NEJM Evidence). However, we found no study assessing whether blood count indices could be used to infer the presence of clonal haematopoiesis.

**Added value of this study:** Here we show that CH driven by mutations in genes associated with high risk of progression to myeloid neoplasia can be reliably differentiated using ML approaches applied on peripheral blood indices; however, low-risk forms of CH (driven by mutations in the *DNMT3A* or *TET2* genes) cannot be reliably inferred from CBC indices. While optimising the model we identified challenges in upscaling its applicability; we propose that the integration of single-cell resolution “raw” blood analyser data might overcome these issues. Previous efforts to enhance the scalability of CH screening focused on reducing DNA sequencing costs. Here, we provide a proof-of-concept that an extensively used clinical test, the CBC, can, using machine learning approaches, predict individuals more likely to harbour high-risk CH, who should be prioritised for genetic testing.

**Implications of all the available evidence:** Our study proposes a model for predicting high-risk CH mutations by applying a Random Forest classifier on CBC indices; this represents an important step towards scalable screening for identifying individuals at high risk of developing myeloid neoplasia in the future. This is an attractive approach, as it relies solely on a routine, inexpensive test. Despite good sensitivity, the low prevalence of high-risk CH leads to a low positive predictive value that precludes the use of the predictive model as a population-wide pre-screening tool. To overcome this, we propose the future integration of raw blood analyser data into models like ours to improve the performance and scalability of this approach.

## Introduction

Haematopoiesis, the formation of the cellular components of blood, occurs continuously throughout life. At steady state, haematopoiesis generates 4-5 × 10^11^ cells per day^1–4^ and this vast output is maintained by a small pool of 50,000-200,000 multipotent haematopoietic stem cells (HSCs)^5^ through a cascade of differentiation and proliferation. Somatic mutations accumulate during life, and though most are inconsequential, some can enhance cellular fitness and are positively selected in physiologically normal tissues^6–8^. Clonal haematopoiesis (CH) is an age-related phenomenon that arises when a HSC acquires a somatic driver mutation (i.e. one that increases its fitness), leading to clonal expansion of the cell and its progeny^9,10^. Large population-based studies revealed that the most commonly mutated genes in CH are involved in epigenetic regulation (*DNMT3A, TET2, ASXL1*), signal transduction (*JAK2, GNB1*), DNA damage response and apoptosis (*TP53, PPM1D*), and splicing (*SF3B1, SRSF2, U2AF1*)^9–14^. The prevalence of CH increases with advancing age to affect at least 20% of those over 70 years, in whom the phenomenon is almost universally detectable when deep sequencing approaches are employed^9–14^.

A hallmark of CH is the associated increased risk of incident myeloid neoplasms (MN), a molecularly heterogenous group of blood cancers that include acute myeloid leukaemia (AML), myelodysplastic syndromes (MDS) and myeloproliferative neoplasms (MPN). The overall rate of progression to MN is low (0.5-1% per annum)^9^, but the risk and nature of malignant progression vary according to the mutant driver gene, the size of the clone, and the selection pressures to which the clone is exposed^15,16^. Recent advances have facilitated the precise estimation of the risk of progression from CH to MN^16,17^, such that individuals at high risk can be identified and prioritised for clinical follow-up. CH may precede the development of MN by years^9–11,15,16,18^, and this provides a window during which high-risk clones could be intercepted and targeted to avert or delay the development of MN.

A key impediment to prospective myeloid cancer prevention programmes is the lack of a scalable test to identify CH. At present, CH is identified by Next Generation Sequencing (NGS) of blood DNA targeted to a panel of genes recurrently mutated in MN. However, NGS is not performed in routine clinical practice and is impractical and costly to perform at scale. An alternative approach is to leverage low-cost, scalable, routine clinical tests to identify individuals likely to harbour CH who can be prioritised for sequencing. The complete blood count (CBC) is an inexpensive, routine clinical test, and CBC indices such as the red cell distribution width (RDW) and mean cell volume (MCV) are associated with progression from CH to MN^18^. We therefore sought to explore whether machine learning (ML) models could predict individuals with CH based on CBC features by analysis of paired CBC and whole exome sequencing (WES) data from 431,531 United Kingdom Biobank (UKB) participants.

## Methods

### Study design and participants

We utilised data from the UKB (https://www.ukbiobank.ac.uk/), a population-based cohort of 502,536 volunteers recruited to the United Kingdom recruited between 2006-2010 and aged between 37 and 73 years at recruitment^19^. Participants’ data was accessed under approved UKB applications number 56844 and 69328.

To derive a dataset for use in our ML pipeline, we excluded UKB participants with any missing CBC variables and those without WES data. Since CH is defined by the presence of a leukaemia-associated somatic driver mutation in an individual without an apparent blood neoplasm, participants with a previous diagnosis of a haematological malignancy were excluded from the final dataset, as were those who developed an incident haematological malignancy within 30 days of recruitment to the UKB. After exclusions, 431,531 participants were retained for downstream analyses.

### Variable selection

We extracted all CBC variables measured in the UKB (n=22), and augmented the feature set with the participants’ age and sex. Some CBC variables are closely related or derived from one another; to assess collinearity we computed a pairwise Spearman’s rank correlation coefficient (r_s_) and excluded variables with a |r_s_| ≥ 0.9. This led us to exclude haematocrit, high light scatter reticulocyte count and the total white blood cell count, whilst retaining their highly correlated counterpart features (haemoglobin concentration, reticulocyte count and neutrophil count, respectively). Nucleated red blood cell count (NRBC) was also excluded as it exhibited near-zero variance (106 unique values, NRBC=0 in 98.9% of UKB participants).

### Identification of clonal haematopoiesis from whole exome sequencing data

CH was identified from whole exome sequencing (WES) of blood DNA from 431,531 UKB participants as previously described^16^ (see Supplement). UKB participants were subsequently labelled as “any-driver-CH” or “no CH” based on the presence or absence of a driver mutation(s) at VAF ≥ 2%. For input to gene-specific models of CH, we additionally labelled UKB participants by driver gene (e.g. “*TET2*-CH”, “*SRSF2*-CH”, etc vs “no CH”). Individuals with ≥2 driver mutations were labelled on the gene with the highest VAF.

### Supervised machine learning model development

Having derived ground truth levels from WES data, ML models were subsequently built for “any-driver-CH” (variant allele frequency, VAF, ≥ 2% with a driver mutation in any CH gene), “large clone any-driver-CH” (as previous but VAF ≥ 10%), and each driver gene CH subtype.

To develop a binary classifier for predicting the presence/absence of CH, we trained and evaluated a selection of tree-based machine learning models: Decision Trees, Random Forests and Extreme Gradient Boosting (XGBoost) Trees. Tree-based approaches were preferred since the set of input features was heterogeneous (continuous and categorical); moreover, these models, augmented with statistical analyses, may also capture the interaction between features. Aside from the assessment of near-zero variance and collinearity, no further pre-processing was applied to the input dataset.

All 18 CBC parameters were used as features, in addition to basic demographic data (age at sampling and sex). Since the UKB CH dataset was imbalanced, with significantly more controls (no CH) than cases (CH), a random down-sampling was performed to achieve a 1:1 ratio of cases:controls in the input data, to enhance model training and convergence; this down-sampling process was repeated ten times iteratively (Supplementary Figure 1). Subsequently, down-sampled datasets were partitioned on 80:20 training:test ratio.

All models were built using ten repeats of ten-fold cross-validation setups; a grid-search approach was used to tune the relevant hyperparameters (Supplementary Table 1). To avoid technical bias from the down-sampling step, a modified cross-validation was applied, training and evaluating each ML model ten times iteratively, each time using a different random down-sample of the majority (control) class, thereby quantifying the robustness and stability of each model to variation in the subset of control samples or train/test partition (Supplementary Figure 1). Model performance was assessed on the unseen test data, on receiver operating characteristic (ROC) curves and area under the curve (AUC), in addition to sensitivity and specificity.

**Table 1:**
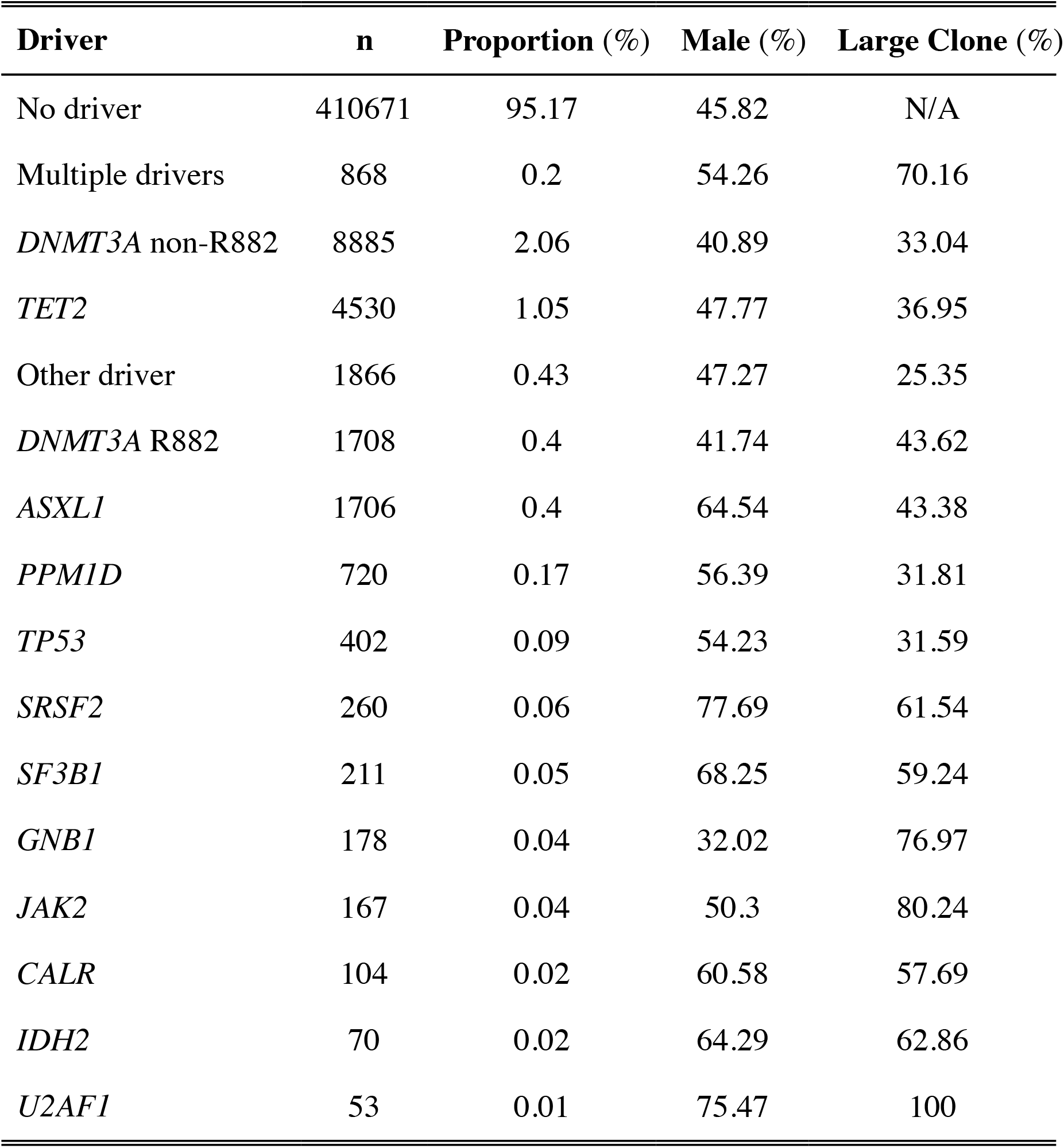
Clonal haematopoiesis proportions in the filtered cohort (n = 431,531).

| Driver | n | Proportion (%) | Male (%) | Large Clone (%) |
| --- | --- | --- | --- | --- |
| No driver | 410671 | 95.17 | 45.82 | N/A |
| Multiple drivers | 868 | 0.2 | 54.26 | 70.16 |
| *DNMT3A* non-R882 | 8885 | 2.06 | 40.89 | 33.04 |
| *TET2* | 4530 | 1.05 | 47.77 | 36.95 |
| Other driver | 1866 | 0.43 | 47.27 | 25.35 |
| *DNMT3A* R882 | 1708 | 0.4 | 41.74 | 43.62 |
| *ASXL1* | 1706 | 0.4 | 64.54 | 43.38 |
| *PPM1D* | 720 | 0.17 | 56.39 | 31.81 |
| *TP53* | 402 | 0.09 | 54.23 | 31.59 |
| *SRSF2* | 260 | 0.06 | 77.69 | 61.54 |
| *SF3B1* | 211 | 0.05 | 68.25 | 59.24 |
| *GNB1* | 178 | 0.04 | 32.02 | 76.97 |
| *JAK2* | 167 | 0.04 | 50.3 | 80.24 |
| *CALR* | 104 | 0.02 | 60.58 | 57.69 |
| *IDH2* | 70 | 0.02 | 64.29 | 62.86 |
| *U2AF1* | 53 | 0.01 | 75.47 | 100 |

From the Random Forests models, we determined variable importance by computing the mean decrease in node impurity from splitting on each feature (measured by Gini index), averaged across all trees and across each of the ten repeats of model-building, using the importance() function from the randomForest package in R (v4.7.1)^21^. The consistency across top-ranked variables per driver was visualised using quantitative Venn diagrams (upset plots, ComplexUpset package) on the top two variables. The feature selection was performed by ranking all *n* features by importance, in descending order, and iteratively excluding the least informative feature, to determine a minimum set of highly predictive features.

To assess the scalability of the final model in a “real-life” setting, i.e. with class imbalance (more controls (no CH) than cases (CH), we added unseen control cases to the test set to match the prevalence of CH cases in the test set to the prevalence of CH cases in the UKB cohort. We examined the trade-off of sensitivity (which is independent of prevalence), positive predictive value (which is dependent on prevalence) and the model prediction score, using this to determine the optimal cut-off score, that minimises the false positives whilst retaining adequate sensitivity.

All ML models were built using the Caret v6.0.91 package in R v3.6.3^20^. A full list of packages used is available in Supplementary Methods. All code used to implement our ML framework is publicly available on GitHub: https://github.com/billydunn/chic.

## Results

After excluding those with missing CBC data (n=32,670), missing WES data (n=36,368), or a prevalent diagnosis of a haematological malignancy (n=1840), CH (VAF ≥2%) was identified in 20,860/431,531 (4.8%) UKB participants, of whom 7637/20,860 (36.6%) had large clone CH (VAF ≥10%; Figure 1,Table 1). Using this UKB dataset, we developed a range of tree-based models using our ML framework, which we henceforth refer to as CHIC (Clonal Haematopoiesis Inference from Counts).

**Figure 1:**
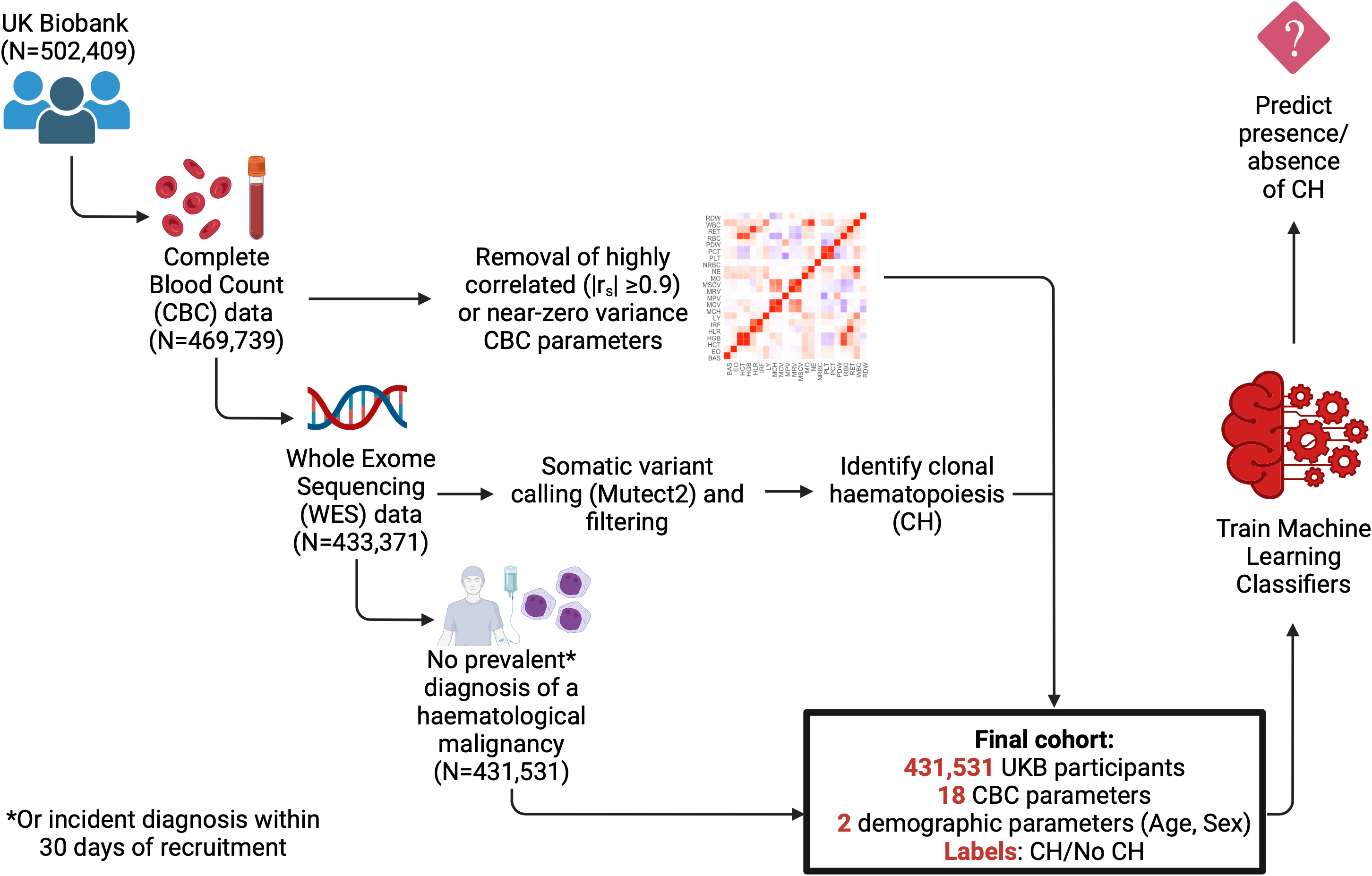
Final cohort derivation in the UK Biobank. We selected only those with all CBC parameters (that is, no missing indices) (n = 469,739). We apply further filtering to include only those who additionally have WES data available, to facilitate identification of CH. Concurrently, we compute Spearman correlation and select only one CBC parameter where two parameters exhibit high positive or negative correlation (|r_s_| ≥0.9). Finally, we exclude individuals who were annotated as having a prevalent diagnosis of a haematological malignancy, or those who had an incident diagnosis shortly after recruitment/blood draw (using an arbitrary threshold of within 30 days of recruitment). This gives us a final dataset of 431,531 participants, each labelled as “CH” or “no CH” for input into our downstream supervised ML pipeline.

We firstly examined whether CH could be predicted from CBC data in the UKB using models agnostic to underlying driver mutations (henceforth “any-driver-CH”). Using CHIC we generated binary classifiers (CH/no CH) of any-driver CH using tree-models with 18 CBC variables augmented with age and sex as features. Classifiers of any-driver CH were better than random, but with limited performance across all model types (median AUC on unseen test set 0.62, 0.64 and 0.62 for DT, RF and XGB models respectively) (Figure 2A).

**Figure 2:**
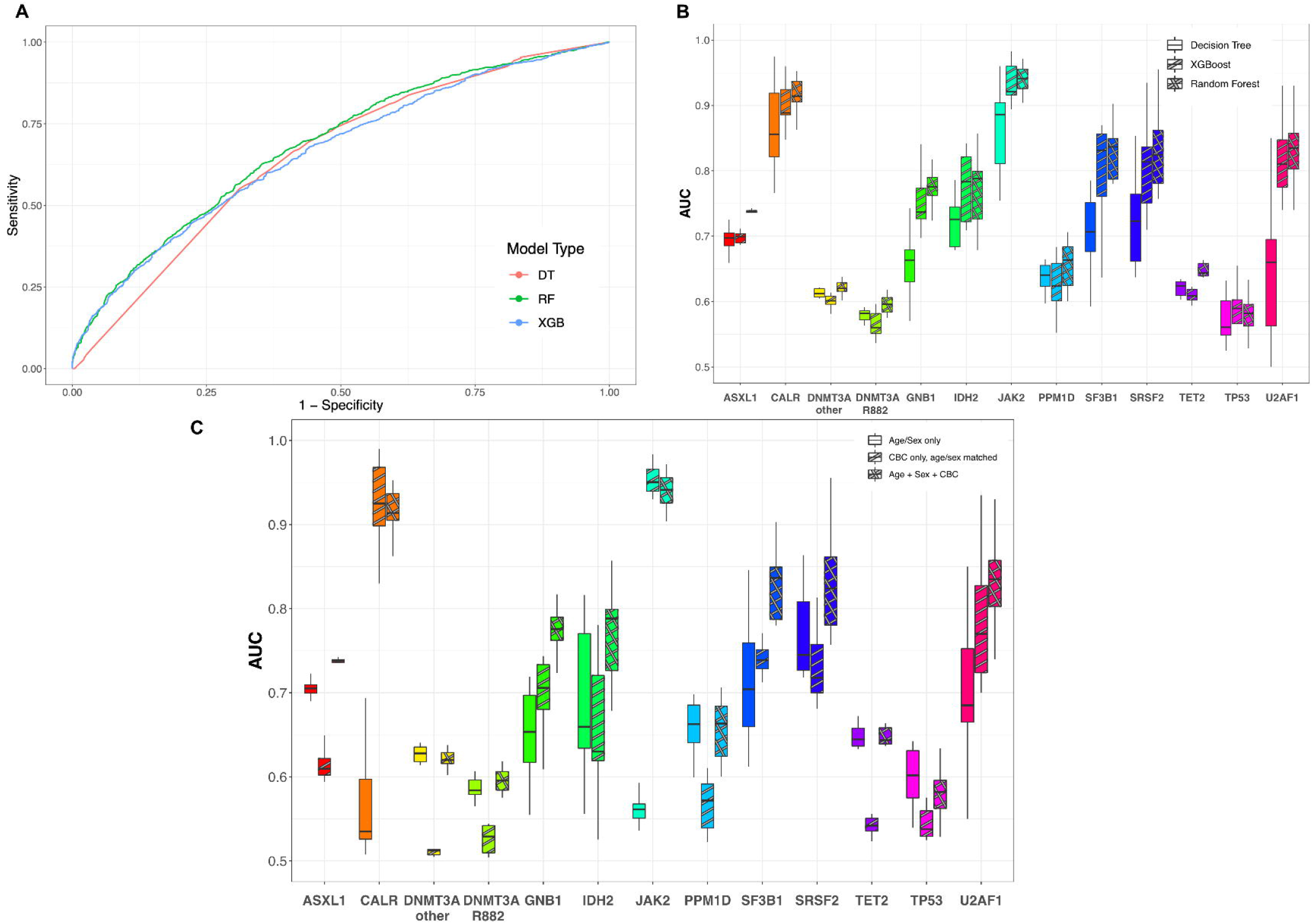
Performance of machine learning classifiers to predict the presence of clonal haematopoiesis. Panel A shows the receiver operating characteristic (ROC) curve for classifiers predicting any-driver CH using age, sex and 18 CBC parameters as features. DT = Decision Tree, RF = Random Forest and XGB = eXtreme Gradient Boosting models. In each case, the ROC curve for the model approximating the median AUC (area under the ROC curve) from ten repeats of model training is shown. Panel B shows the performance of driver gene-specific models across all three model types (DT, RF, XGB), using the same features. Boxplots are derived from the ten repeats of model building; whiskers show the range of AUC values. Panel C shows the performance of RF classifiers of driver gene-specific CH, using either: age and sex as the only features; or CBC features only (with age and sex matching of cases to controls, to capture the predictive performance of CBC indices alone); or age, sex and CBC features (without age and sex matching, thereby capturing the predictive performance of CBC indices in combination with basic demographics).

CH is a molecularly heterogenous entity, and we posited that the nature and strength of the CBC phenotype conferred by a somatic mutation may vary according to the specific driver gene. We trained driver gene-specific binary classifiers (with labels driver gene CH/no CH) using the same input variables as for the any-driver CH models. The most prevalent forms of CH, driven by mutations in *DNMT3A* and *TET2*, were not robustly detectable; this conclusion held for *DNMT3A*-R882 hotspot mutations, which are associated with a slightly higher risk of transformation to AML^16^ (median AUC 0.60, 0.62 and 0.64 for *DNMT3A*-R882, *DNMT3A*-non R882 and *TET2* RF models respectively) (Figure 2B). By contrast, CH driven by lower prevalence but higher risk driver mutations in the genes *JAK2, CALR, SF3B1, SRSF2* and *U2AF1* performed well (median AUC 0.94, 0.91, 0.84, 0.82, 0.84 respectively for RF models) (Figure 2B). Since Random Forests (RF) models generally exhibited the best performance across the driver genes (Figure 2B, Supplementary Table 2), we focused on further developing and exploring RF models.

**Table 2:** Confusion matrix for predictions made by the classifier of high-risk CH on the unseen test set (n=86,306), using the previously determined stringent cutoff to determine whether the classifier predicts the positive class (high-risk CH) or the negative class (no high-risk CH).

|  |  |  |  |
| --- | --- | --- | --- |
| Reference | High-risk CH | 32 | 127 |
|  | No high-risk CH | 365 | 85782 |
|  |  | High-risk CH | No high-risk CH |
|  |  | Prediction |  |

CH is strongly associated with age, whilst some driver genes exhibit sex bias. To understand the influence of age and sex in the RF models, we trained each set of driver gene-specific RF models in three iterations: i) with age and sex as the only features, ii) with CBC indices as the features, whilst age- and sex-matching cases to controls (to capture the predictive performance of CBC alone), and iii) with age, sex and CBC indices as features, without age-and sex-matching of cases/controls (to capture the predictive performance of both basic demographics and CBC indices). The performance of models trained with only age and sex as features was generally poor (median AUC <0.75 in all cases, Figure 2C); an exception was the age/sex-only model of *SRSF2-*CH, in line with the sharp rise in prevalence of *SRSF2*-CH with advancing age and its strong association with male sex^15^.

Classifiers of CH driven by high-risk genes *JAK2, CALR, SF3B1, SRSF2* and *U2AF1* also performed best when using CBC indices as features and age/sex matching cases to controls in the training and test sets. The predictability of the presence of CH driven by mutations in splicing factor genes (*SF3B1, SRSF2* and *U2AF1*) was augmented when age and sex were added as features and age/sex matching was omitted. Acknowledging the age and sex predictive power, we added these features to CBC indices in subsequent models.

Since CH with mutations in any of *JAK2, CALR, SF3B1, SRSF2* or *U2AF1* was more predictable from CBC indices and more clinically relevant (associated with high risk of progression to MN), we next combined all predictors into a single binary classifier of “high-risk CH”, to predict the presence/absence of a mutation in any of these five genes (training on input data labelled as “high-risk CH” vs “no high-risk CH”). The resulting median AUC was 0.85 on the unseen test set, Figure 3A); the model also predicted the presence of large (VAF ≥10%) high-risk clones (median AUC on unseen test set 0.90, Supplementary Figure 2).

**Figure 3:**
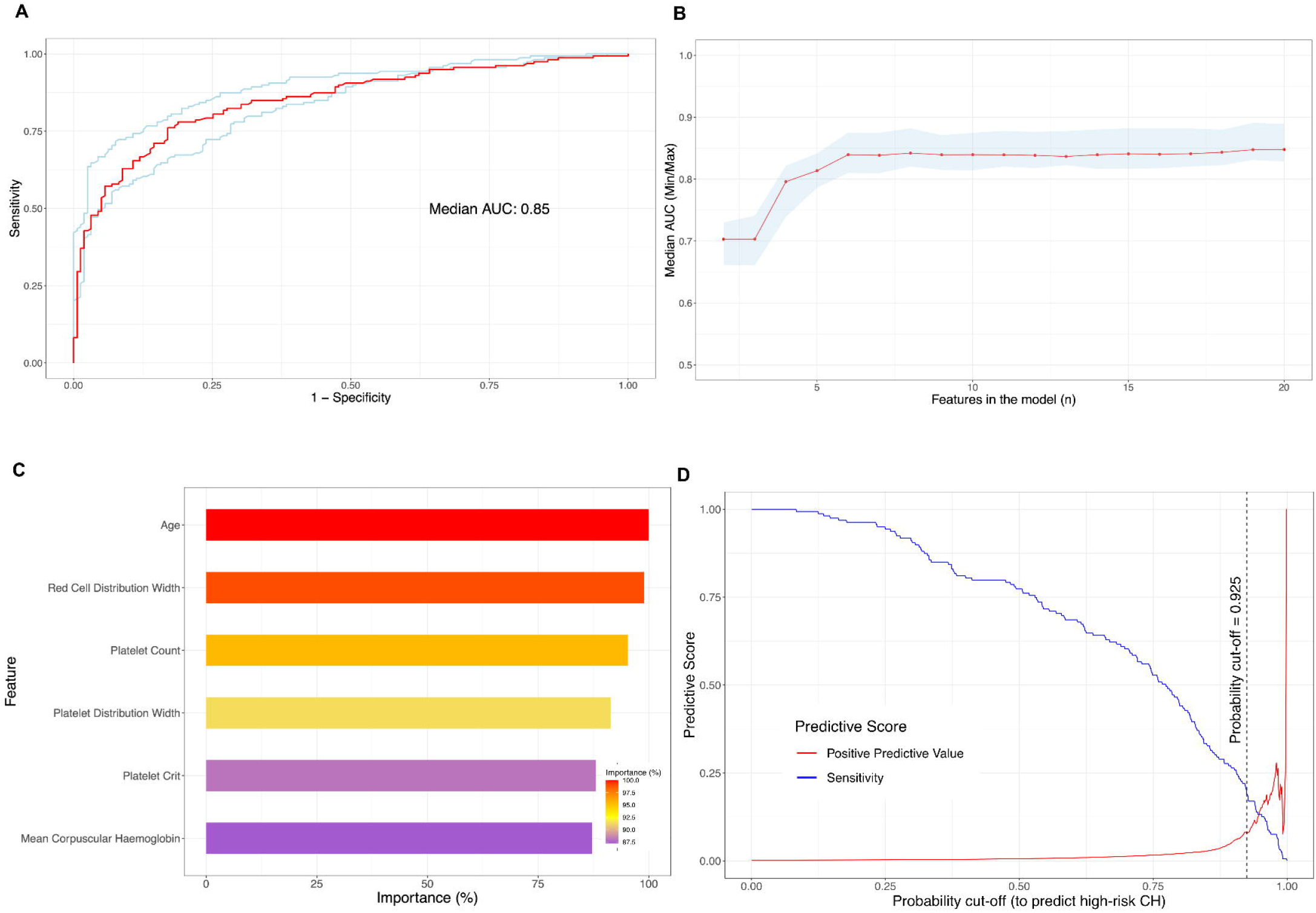
Optimisation, Variable Importance and Performance of a classifier of high-risk CH. Panel A shows the ROC curve for this RF model, which has been constructed and AUC calculated based on performance in the unseen test set. Red, performance of model approximating the median AUC. Upper and lower bounds represent performance of the models with the maximum and minimum AUC from ten repeats of model training respectively. Panel B shows the impact of iterative feature selection on model performance (by AUC), demonstrating that performance is stable with only six features. Panel C shows variable importance (by Gini Index, scaled to the most important variable) of features in our six-feature classifier. Panel D shows the trade-off between sensitivity (blue) and positive predictive value (red) for this six-feature classifier.

To further refine the classifier of high-risk CH with VAF ≥2%, we performed iterative feature selection, incrementally excluding the least discriminative feature, to obtain the minimal stable set of highly discriminative features; this demonstrated that our classifier of high-risk CH had undiminished performance using only six features: age at blood sampling, red cell distribution width (RDW), platelet count, platelet distribution width (PDW), platelet crit and mean corpuscular haemoglobin (MCH) (Figure 3B-C). We therefore chose this compact high-risk CH model to explore further, selecting the model that most closely approximated the median AUC across the ten models built using our iterative pipeline.

Next, we assessed the optimal prediction score cut-off (threshold) for our compact high-risk CH model by examining the trade-off between sensitivity and positive predictive value (PPV) (Figure 3D). In our UKB cohort, high-risk CH was rare (795/431,531 UKB participants, prevalence 0.18%): since the PPV is strongly influenced by the prevalence of positive cases, this necessitated the use of a stringent prediction score cut-off to minimise the number of false positives. To achieve this, we chose a cut-off probability of 0.925, giving a PPV of 8.1% and sensitivity of 20.1% in our unseen test cohort (n=86,306), whilst maintaining the specificity and negative predictive value (NPV) of >99.5% (Table 2).

A key limitation of the UKB is the low WES coverage, with the driver genes *JAK2, SF3B1* and *U2AF1* all having a median coverage of ≤ 31 reads^16^, rendering variant calling insensitive to smaller clones. As such, we examined outcomes for the 365 “false positive” cases identified by our high-risk CH classifier, and found that 38/365 (10.4%) developed MN at a median of 5.2 years from sampling. By contrast, only 317/85,782 (0.4%) percent of “true negatives” developed MN. Since CH is the shared precursor of the vast majority of MNs, these observations strongly suggest that the “false positive” individuals had CH below the limit of detection of WES.

To further explore this hypothesis, we searched for low VAF hotspot mutations amongst 38 individuals who developed MPN, but were not found to have this hotspot mutation by standard variant calling. To do so, we used “pileup” to detect hotspot mutant reads that were filtered out by the stringent criteria of standard calling; this revealed that 13/38 of apparently false positives who developed incident MN had detectable CH mutations by this method, including 11 with driver mutations in *JAK2*, a low coverage gene. This strongly suggests that we underestimated our model performance due to the constraints of WES.

Further examination of cases identified by CHIC revealed an enrichment in cases with thrombocytosis, suggestive of undiagnosed or unannotated MPN rather than CH (Supplementary Figure 3). Similarly, a few cases had cytopenias that would fall into the diagnostic criteria for CCUS (clonal cytopenia of undetermined significance) or MDS^22^. To overcome this, we constrained our training/test sets to individuals without cytopenias, thrombocytosis or erythrocytosis, (see Supplementary Methods) and retrained our high-risk classifier. This led to only a minor reduction in performance (median AUC on unseen test set 0.80, Supplementary Figure 4), however, this exacerbated the trade-off between sensitivity and PPV, leading to sensitivity and PPV of only 11.3% and 2.0% respectively at our proposed cutoff probability of 0.875 (Supplementary Figure 4).

**Figure 4:**
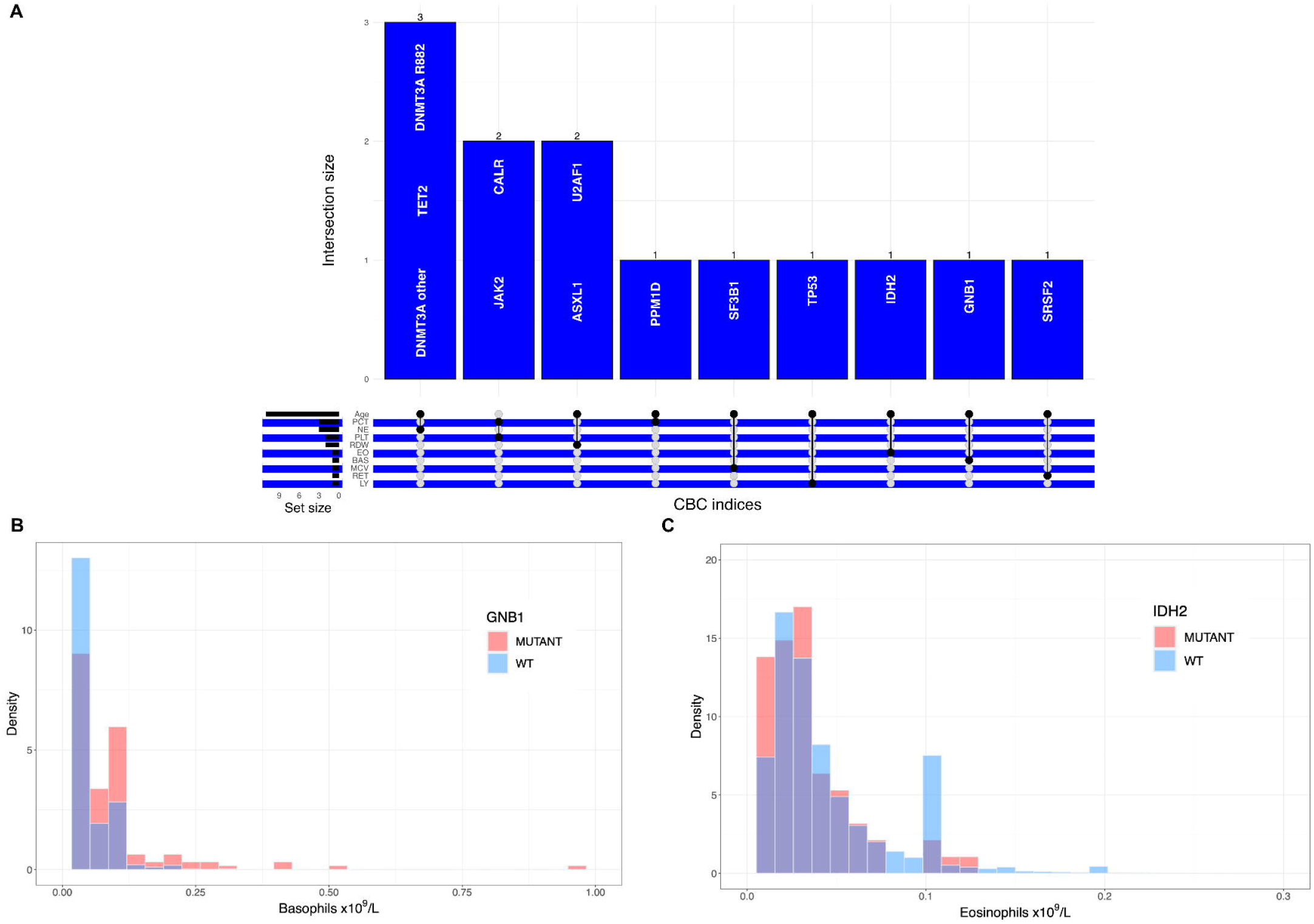
Machine learning models of driver gene CH for biological inference. Panel A shows an Upset plot generated by computing variable importance and summarising the overlap (vertical bars) between the top two most important variables in RF classifiers of driver gene CH. This captures expected associations (*JAK2* and *CALR* share platelet crit and platelet count as their top two variables), but also unveils unexpected associations, such as the importance of basophil count for predicting the presence of *GNB1*-CH and the importance of eosinophil count in predicting the presence of *IDH2*-CH. PCT = platelet crit, NE = neutrophil count, PLT = platelet count, RDW = red cell distribution width, EO = eosinophil count, BAS = basophil count, MCV = mean cell volume, RET = reticulocyte count, LY = lymphocyte count. Panel B shows a histogram of basophil counts in carriers of *GNB1*-CH (n = 178) versus those without (n = 431,353); the basophil count is shifted to the right in those with *GNB1*-CH, who have a relatively high prevalence of basophilia. Panel C shows the histogram of eosinophil counts in individuals with (n = 70) and without (n = 431,461) *IDH2*-CH; there is a higher proportion of absolute eosinopenia (i.e. eosinophil count = 0) in individuals with *IDH2*-CH. In both panels B and C, the y axis is density, to facilitate direct comparison between imbalanced classes.

In addition to their use for prediction, CHIC ML models could also uncover novel associations between driver mutations and CBC indices. By evaluating variable importance across all the driver-gene-specific classifiers, and summarising the overlap between the top two features in each model (Figure 4A), we observed known or expected associations: age was highly predictive across models, *JAK2*-CH and *CALR*-CH shared platelet count and platelet crit as important features whilst MCV was predictive of *SF3B1*-CH. Unexpected associations were also revealed: for example, the basophil count was discriminative for predicting the presence *GNB1*-CH only, whilst eosinophil count was discriminative for the presence of *IDH2*-CH. Examining the distribution of each of these CBC variables in the UKB, we found that individuals with *GNB1*-CH had a significantly increased basophil count (p = 5.93 × 10^−11^, Wilcoxon Rank Sum test), with a 4.5-fold increase in the prevalence of basophilia >0.1 × 10^9^/L and 8.6-fold increase in the prevalence of basophilia >0.2 × 10^9^/L, relative to participants without a *GNB1* driver mutation (13.8% vs 3.2% and 5.0% vs 0.6% for basophilia >0.1 and >0.2 × 10^9^/L, n = 178/431,353 for *GNB1* mutant/wild-type respectively) (Figure 4B). Individuals with *IDH2*-CH had significantly lower eosinophil counts (p = 3.63 × 10^−10^, Wilcoxon Rank Sum test) and a propensity to eosinopenia, with 12/92 (13.0%) participants with *IDH2*-CH having absolute eosinopenia (eosinophils = 0 × 10^9^/L) and 45/92 (48.9%) having an eosinophil count <0.1 × 10^9^/L, by contrast individuals without *IDH2* mutations had rates of eosinopenia of 2.9%/20.8% for absolute/<0.1 eosinopenia respectively (n = 70/431,461 for *IDH2* mutant/wild-type respectively) (Figure 4C). Only *IDH2*-CH demonstrated a significant association between eosinophil count and clone size (r_s_ = -0.51, p = 2.67 × 10^−7^); we observed no such association between *GNB1*-CH and basophil count (r_s_ = 0.13, p = 0.09) (Supplementary Figure 5), though of note basophils are the rarest of the white blood cell subsets and as a result their counts are zero-biased, which may have confounded any putative association.

## Discussion

We developed the CHIC framework and assessed a RF classifier that predicts the presence of high-risk CH from just five CBC variables and an individual’s age. This approach, named Clonal Haematopoiesis Inference from Counts (CHIC), can discriminate between individuals with and without mutations in five CH genes associated with high-risk of developing MN. Notably, CHIC retained an ability to discriminate high-risk CH cases from controls even amongst individuals without cytopenias, erythrocytosis or thrombocytosis, suggesting it may highlight individuals that may not otherwise come to medical attention. CHIC is an important first step towards developing a scalable screening test to identify individuals likely to harbour high-risk CH, who would then be prioritised for targeted NGS. This would not only vastly reduce the number needed to screen (NNS) per case of high-risk CH identified, but it would also justify the need to perform genetic testing. Even with its current limitations, the use of CHIC with a stringent cut-off probability on individuals without cytopenia or thrombo-/erythrocytosis would still markedly reduce the NNS from 727 to 40 individuals per case of high-risk CH (based on the prevalence of high-risk CH in an unselected population vs in those predicted as having high-risk CH by CHIC). The implementation of a scalable screening test would represent a significant milestone in myeloid cancer prevention, by addressing a key bottleneck in recruitment to interventional studies.

However, despite its promising metrics as a screening test, the performance of CHIC in an unselected population was limited by the rarity of high-risk CH, necessitating ceding sensitivity to achieve an acceptable PPV. Performance was further reduced for the restricted analysis of individuals without cytopenias or thrombo-/erythrocytosis. By re-training our model in this population, we found that CHIC was still able to discriminate individuals with and without high-risk CH, but the resultant small reduction in AUC (0.80 vs 0.85) exacerbated the difficulty in balancing sensitivity and PPV, precluding its use at population scale.

One approach for enhancing the performance of CHIC is to target its use on a population with a higher prevalence of high-risk CH. CHIC was trained within the age constraints of the UKB, but since the prevalence of high-risk mutations in splicing factors (*SF3B1, SRSF2, U2AF1*) rises sharply over the age of 70 years, we anticipate that application of CHIC in an older population would result in improved performance. Similarly, targeting CHIC to individuals with a polygenic^15,23^ or monogenic^24^ predisposition to CH is also likely to improve its performance/PPV.

An alternative approach would be to integrate higher-resolution CBC data into the CHIC classifier, to improve its ability to identify high-risk CH. Some of the most discriminative CBC indices for high-risk CH are derived summary stastics e.g. RDW, PDW and MCH calculated from single-cell measurements (i.e. RDW is a measure of variation in red cell volumes). The integration of the raw or otherwise summarised single-cell measurements has the potential to improve the prediction of high-risk CH, for example by revealing a fraction of cells with distinct indices arising from the CH clone or identifying other characteristic patterns of variation in these measurements; such raw (or “non-classical”) CBC traits have recently been exploited to explore genetic associations with blood cell morphology^25^.

Beyond MN prevention, CH is of wider public health relevance due to its association with non-haematological disorders, most notably atherosclerotic heart disease. Since *JAK2*-CH exhibits the strongest association with cardiovascular outcomes^26^, and was also the most amenable to prediction in our study, we anticipate that CHIC may also have utility in the primary prevention of cardiovascular disease by facilitating the identification of individuals with *JAK2*-CH. By retrofitting CH screening on to a routine blood test, we believe our CHIC approach presents an important step towards scalable, practical and inexpensive ML-based screening for high-risk CH and provides a proof-of-concept that individuals with high-risk CH can be differentiated from those without, based on CBC indices.

## Data sharing

All data used in this study are publicly available from the UK Biobank (https://www.ukbiobank.ac.uk/). Researchers may apply for access to the UK Biobank data via the Access Management System (https://www.ukbiobank.ac.uk/enable-your-research/apply-for-access).

## Code availability

Scripts used to query the UK Biobank dataset are available from: https://github.com/IsabellaWithnell/Predicting_CH. Scripts used to implement the machine learning framework described in the manuscript are available from: https://github.com/billydunn/chic.

## References

1. Harker, L. A. & Finch, C. A. Thrombokinetics in man. J Clin Invest 48, 963–974 (1969).

2. Kaushansky Kenneth. Lineage-Specific Hematopoietic Growth Factors. New England Journal of Medicine 354, 2034–2045 (2006).

3. Cosgrove, J., Hustin, L. S. P., de Boer, R. J. & Perié, L. Hematopoiesis in numbers. Trends Immunol 42, 1100–1112 (2021).

4. Sender, R. & Milo, R. The distribution of cellular turnover in the human body. Nat Med 27, 45–48 (2021).

5. Mitchell, E. et al. Clonal dynamics of haematopoiesis across the human lifespan. Nature 606, 343–350 (2022).

6. Martincorena, I. et al. High burden and pervasive positive selection of somatic mutations in normal human skin. Science 348, 880–886 (2015).

7. Martincorena, I. et al. Somatic mutant clones colonize the human esophagus with age. Science 362, 911–917 (2018).

8. Lee-Six, H. et al. Population dynamics of normal human blood inferred from somatic mutations. Nature 561, 473–478 (2018).

9. Jaiswal, S. et al. Age-related clonal hematopoiesis associated with adverse outcomes. N Engl J Med 371, 2488–2498 (2014).

10. Genovese, G. et al. Clonal hematopoiesis and blood-cancer risk inferred from blood DNA sequence. N Engl J Med 371, 2477–2487 (2014).

11. Xie, M. et al. Age-related mutations associated with clonal hematopoietic expansion and malignancies. Nat Med 20, 1472–1478 (2014).

12. McKerrell, T. et al. Leukemia-associated somatic mutations drive distinct patterns of age-related clonal hemopoiesis. Cell Rep 10, 1239–1245 (2015).

13. Young, A. L., Challen, G. A., Birmann, B. M. & Druley, T. E. Clonal haematopoiesis harbouring AML-associated mutations is ubiquitous in healthy adults. Nat Commun 7, 12484 (2016).

14. Zink, F. et al. Clonal hematopoiesis, with and without candidate driver mutations, is common in the elderly. Blood 130, 742–752 (2017).

15. Kar, S. P. et al. Genome-wide analyses of 200,453 individuals yield new insights into the causes and consequences of clonal hematopoiesis. Nat Genet 54, 1155–1166 (2022).

16. Gu, M. et al. Multiparameter prediction of myeloid neoplasia risk. Nat Genet 55, 1523–1530 (2023).

17. Weeks, L. D. et al. Prediction of Risk for Myeloid Malignancy in Clonal Hematopoiesis. NEJM Evidence 2, (2023).

18. Abelson, S. et al. Prediction of acute myeloid leukaemia risk in healthy individuals. Nature 559, 400–404 (2018).

19. Sudlow, C. et al. UK biobank: an open access resource for identifying the causes of a wide range of complex diseases of middle and old age. PLoS Med 12, e1001779 (2015).

20. Kuhn, M. Building Predictive Models in R Using the caret Package. Journal of Statistical Software 28, 1–26 (2008).

21. Liaw, A. & Wiener, M. Classification and Regression by RandomForest. Forest 23, (2001).

22. Khoury, J. D. et al. The 5th edition of the World Health Organization Classification of Haematolymphoid Tumours: Myeloid and Histiocytic/Dendritic Neoplasms. Leukemia 36, 1703–1719 (2022).

23. Kessler, M. D. et al. Common and rare variant associations with clonal haematopoiesis phenotypes. Nature 612, 301–309 (2022).

24. DeBoy, E. A. et al. Familial Clonal Hematopoiesis in a Long Telomere Syndrome. N Engl J Med 388, 2422–2433 (2023).

25. Akbari, P. et al. A genome-wide association study of blood cell morphology identifies cellular proteins implicated in disease aetiology. Nat Commun 14, 5023 (2023).

26. Jaiswal, S. et al. Clonal Hematopoiesis and Risk of Atherosclerotic Cardiovascular Disease. N Engl J Med 377, 111–121 (2017).

