## Supplementary Materials for "Machine learning framework for predicting the presence of high-risk clonal haematopoiesis using complete blood count data: a population-based study of 431,531 UK Biobank participants"

**Table of Contents**


| | |
|---|---|
| Supplementary Methods | Page 2 |
| Supplementary Figures | Pages 3-7 |
| Supplementary Tables | Pages 8-11 |
| Supplementary Files | Page 12 |

### Supplementary Methods

#### Identification of clonal haematopoiesis from whole exome sequencing data

To identify UK Biobank participants with CH driver mutations, we used mutation calls from blood WES data that were previously published by Gu et al[1]. Briefly, Mutect2 was run in tumour-only mode against a panel of 38 CH-associated genes. Germline variants were filtered using a panel of normals. Putative somatic variants were further filtered using FilterMutectCalls, while variants flagged as "germline" were rescued if they were present at least five times in the set of putative somatic variants. To derive a final list of driver mutations, variants were firstly filtered based on number of alternate reads (≥2), presence on forward and reverse strands, minimum read depth (≥7/≥10 for SNVs/indels respectively) and minor allele frequency (<0.001) in gnomAD. The criteria described by Vlasschaert et al.[2] were then used to further filter germline variants and sequencing artefacts. To overcome previously reported issues with mapping at the *U2AF1* locus[3], *U2AF1* mutations were called separately using Samtools[4] mpileup to identify single nucleotide variants (SNVs) at known hotspots, with variants supported by ≥3 reads and at VAF >0.1 retained for downstream analyses.

#### Developing a model of high-risk CH with "normal" CBC indices

To investigate whether our models could detect individuals with CHIP, who, by definition, have relatively normal CBC indices, we further constrained our training and test cohorts to include only UKB participants who did not have a cytopenia (haemoglobin <12/13 g/dL for males/females respectively, neutrophils <1.8 x 109/L, platelets <150 x109/L), erythrocytosis (haemoglobin >16.5/16 g/dL or haematocrit percentage >49/48% for males/females respectively) or thrombocytosis (platelets > 450 x 109/L). These definitions were derived from the definitions of cytopenias, erythrocytosis and thrombocytosis used in the diagnostic criteria for CCUS, MDS and MPN in the 5th edition of the World Health Organisation Classification of Haematolymphoid Tumours. We then used this constrained population as input to our ML model training & validation pipeline, as outlined in our Methods section in the main manuscript.

**Supplementary Figures**

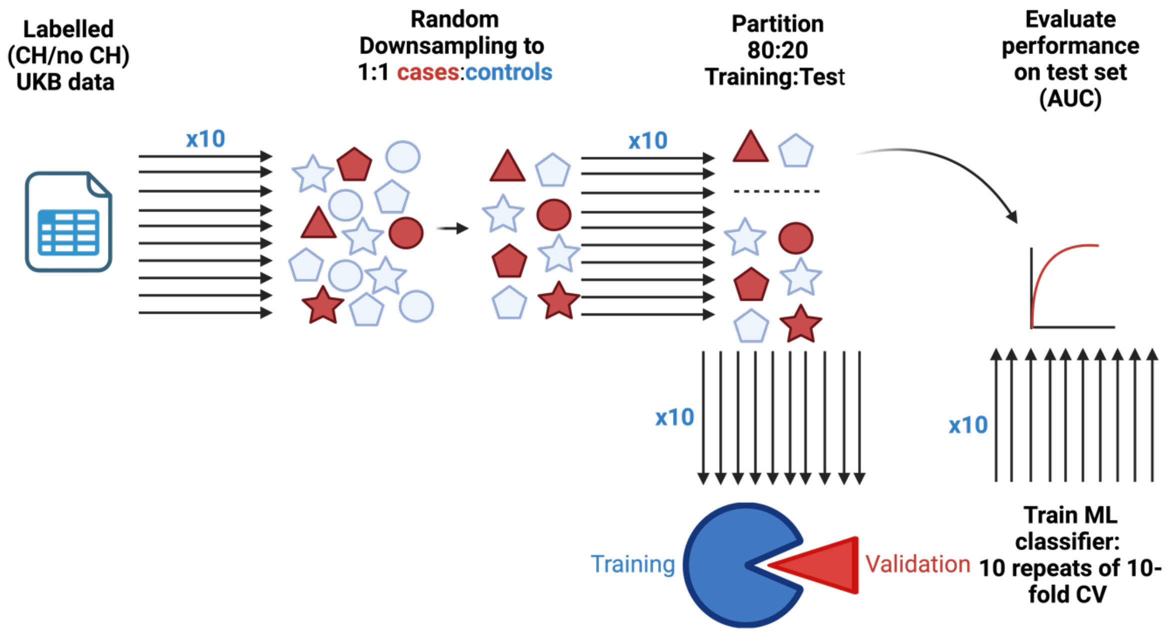


***Supplementary Figure 1****: Overview of our machine learning pipeline. We draw from our master dataset of 431,531 UKB participants with variables age, sex and CBC parameters, each labelled as "CH" or "no CH" (or for gene-specific models, gene-specific CH or no-CH). To enhance model training and convergence, we randomly downsample from this master dataset ten times, to produce ten different datasets each with a 1:1 ratio of cases (CH, red) to controls (no CH, blue). Each dataset has the same set of cases, but a random sample of controls. We then partition the ten datasets in an 80:20 ratio to training:test cohorts, and train ten ML models, each time using ten repeats of ten-fold cross-validation to control for overfitting. Grid search was used to tune the relevant hyperparameters for each model type. The performance of each model was evaluated on the unseen test set, using area under the ROC curve (AUC) as the primary performance measure. By generating the starting dataset ten times, and independently partitioning each dataset and training/evaluating each model, we assess the robustness and stability of each model to variations in the train/test split and in the random sampling of controls.*

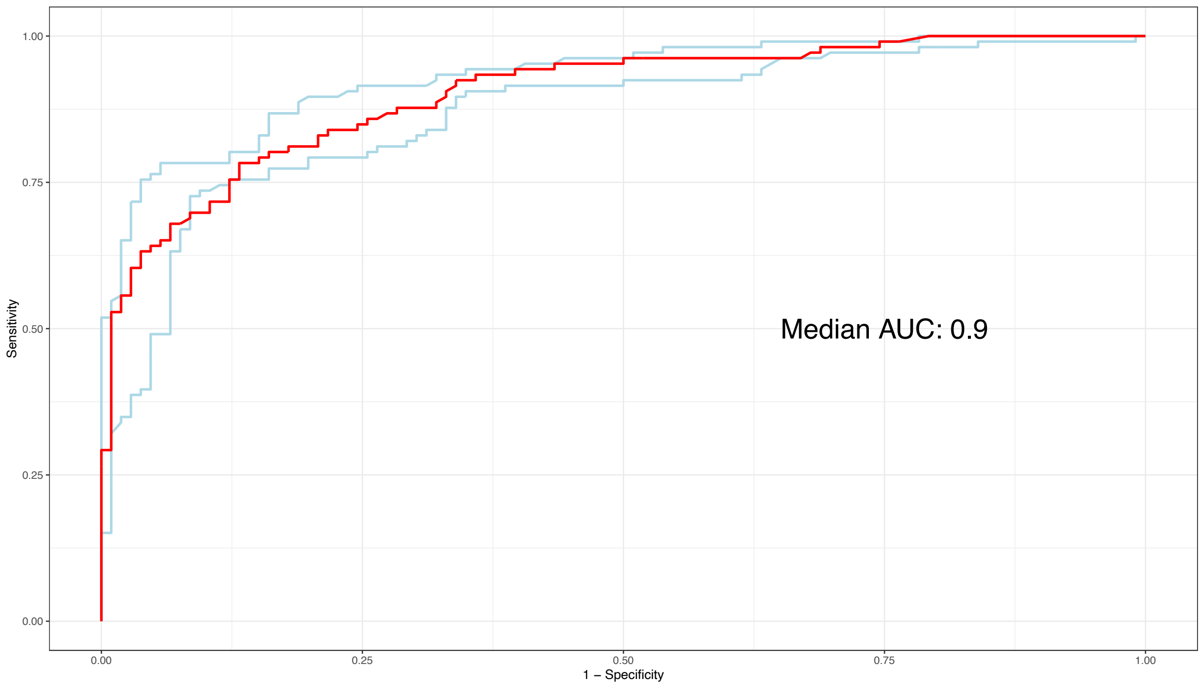


***Supplementary Figure 2****: ROC curve for model of large clone (VAF ≥10%) high-risk CH driven by JAK2, CALR, SF3B1, SRSF2 or U2AF1. This model utilises all 18 CBC variables, age and sex as input features. The ROC curve has been constructed and AUC calculated based on performance in the unseen test set. Red, performance of model approximating the median AUC. Upper and lower bounds represent performance of the models with the maximum and minimum AUC from ten repeats of model training respectively.*

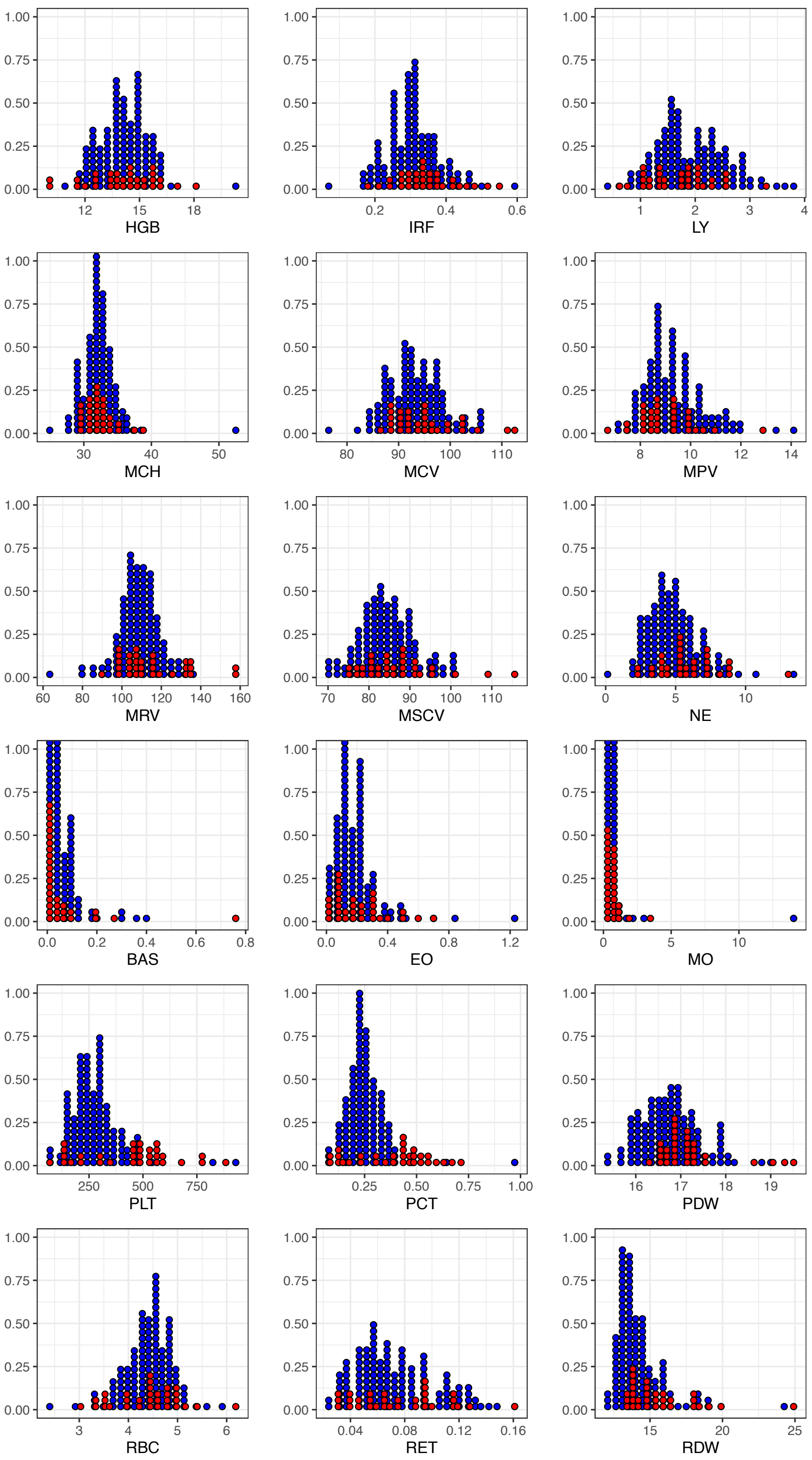


***Supplementary Figure 3:*** *Distribution of CBC indices in high-risk cases in the test cohort. Red = predicted as a case (true positive), blue = predicted as control (false negative), using our proposed stringent cutoff for our six-feature model (input features age, RDW, PLT, PDW, PCT, MCH). Most true positives have high platelet counts, which may be consistent with an unannotated MPN.*

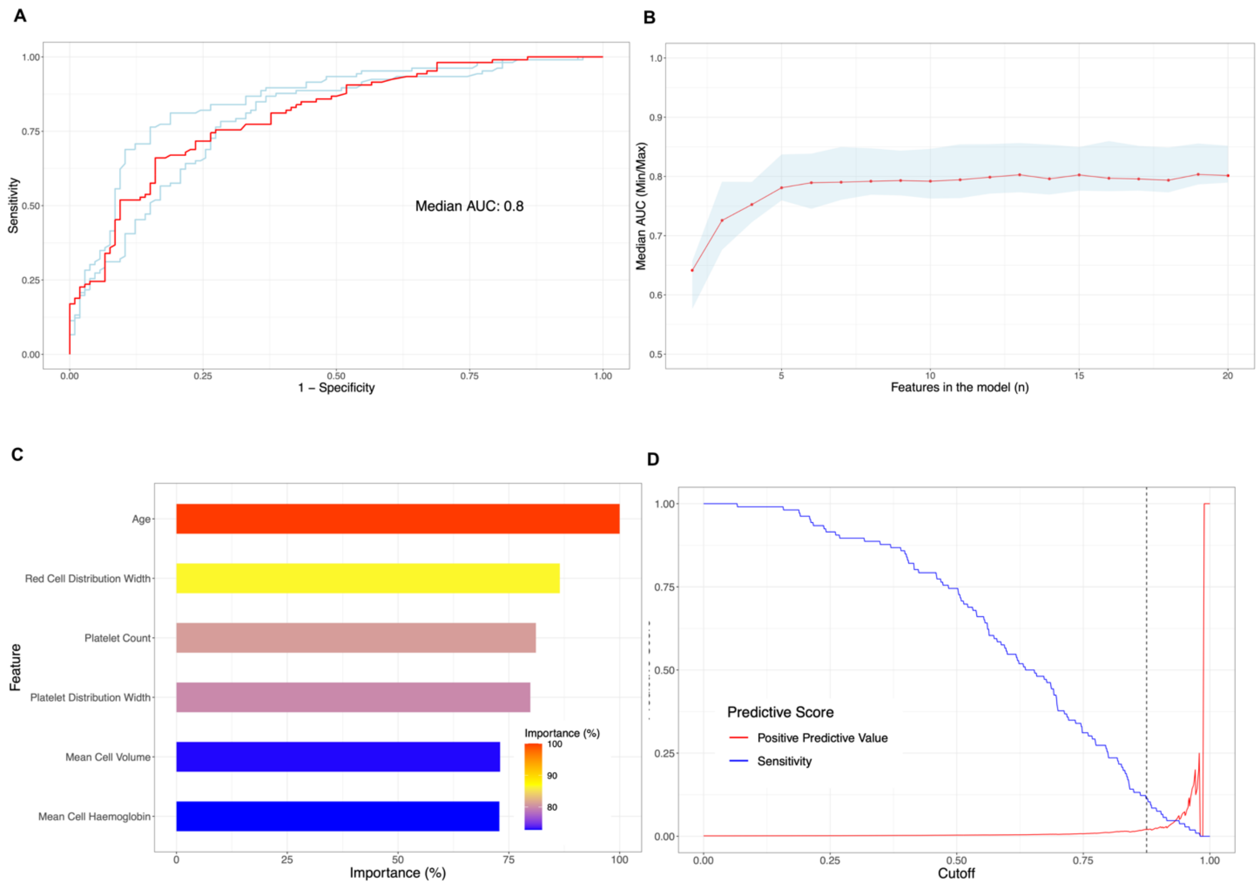


***Supplementary Figure 4****: Optimisation, Variable Importance and Performance of a classifier of High-Risk CH with normal CBC indices. Panel A shows the ROC curve for this Random Forest model, which has been constructed and AUC calculated based on performance in the unseen test set. Red, performance of model approximating the median AUC. Upper and lower bounds represent performance of the models with the maximum and minimum AUC from ten repeats of model training respectively. Panel B shows the impact of iterative feature selection on model performance (by AUC), demonstrating that performance is stable with only six input features. Panel C shows variable importance (by Gini Index, scaled to the most important variable) of features in our six-feature classifier. Panel D shows the trade-off between sensitivity (blue) and positive predictive value (red) for this six-feature classifier.*

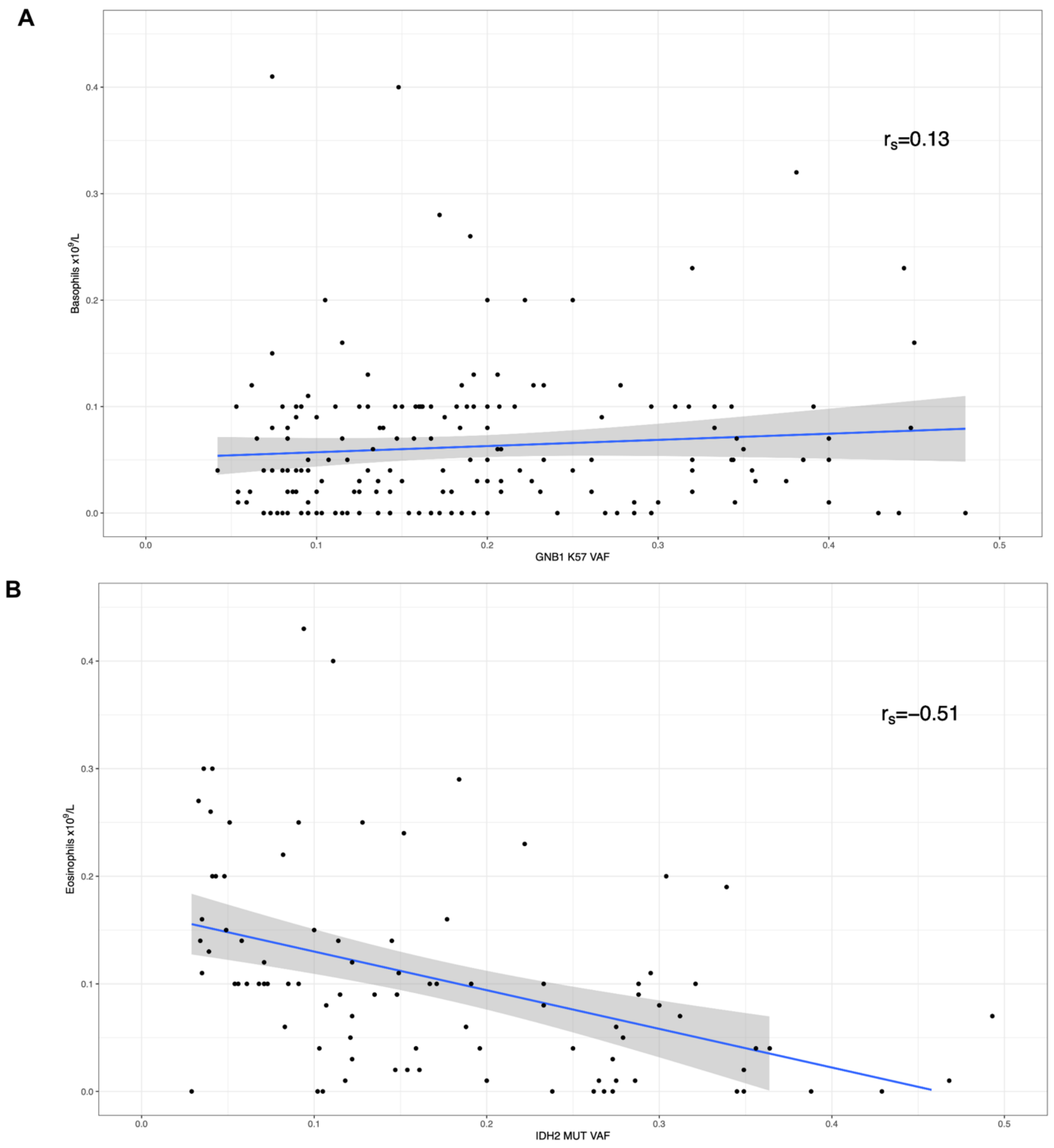


***Supplementary Figure 5:*** *Correlation between basophil/eosinophil count and GNB1/IDH2 mutation variant allele fraction (VAF), respectively. $R_s$ denotes Spearman's rho value. IDH2 clone size exhibits a significant inverse correlation with eosinophil count ($p = 2.67 \times 10^{-7}$), but we observed no significant correlation ($p = 0.09$) between GNB1 clone size and basophil count.*

**Supplementary Tables**

| *Model type* | *Hyperparameters* | *Tuning values* |
|---|---|---|
| DT | cp | 0.001, 0.005, 0.01, 0.15, 0.2, 0.3, 0.4, 0.5 |
| RF | mtry | 2, 4, 6, 8, 10 |
| RF | ntree | 1000 |
| XGB | nrounds | 1000 |
| XGB | eta | 0.3 |
| XGB | max_depth | 2, 4, 6, 8, 10 |
| XGB | gamma | 0 |
| XGB | colsample_by_tree | 1 |
| XGB | min_child_weight | 1 |
| XGB | subsample | 1 |

***Supplementary Table 1:*** *Hyperparameter values tuned using grid search, during model optimisation.*

| *Gene* | *DT* | *RF* | *XGB* |
|---|---|---|---|
| ASXL1 | 0.70 | 0.74 | 0.70 |
| BCOR | 0.50 | 0.53 | 0.53 |
| BCORL1 | 0.50 | 0.52 | 0.54 |
| CALR | 0.86 | 0.91 | 0.89 |
| CBL | 0.54 | 0.52 | 0.59 |
| DNMT3A R882 | 0.58 | 0.60 | 0.56 |
| DNMT3A non-R882 | 0.61 | 0.62 | 0.60 |
| EZH2 | 0.53 | 0.56 | 0.57 |
| GNAS | 0.61 | 0.66 | 0.63 |
| GNB1 | 0.66 | 0.78 | 0.74 |
| IDH2 | 0.73 | 0.79 | 0.78 |
| JAK2 | 0.89 | 0.94 | 0.92 |
| KDM6A | 0.50 | 0.54 | 0.53 |
| PHF6 | 0.50 | 0.57 | 0.57 |
| PPM1D | 0.64 | 0.66 | 0.62 |
| RAD21 | 0.50 | 0.61 | 0.57 |
| RUNX1 | 0.55 | 0.58 | 0.55 |
| SF3B1 | 0.71 | 0.84 | 0.83 |
| SMC3 | 0.50 | 0.57 | 0.55 |
| SRSF2 | 0.72 | 0.82 | 0.81 |
| STAG2 | 0.52 | 0.52 | 0.54 |
| TET2 | 0.62 | 0.64 | 0.61 |
| TP53 | 0.56 | 0.58 | 0.58 |
| U2AF1 | 0.66 | 0.84 | 0.81 |
| ZRSR2 | 0.55 | 0.57 | 0.58 |
| Any-driver CH | 0.62 | 0.64 | 0.62 |

***Supplementary Table 2:*** *Performance across model types and driver genes. DT, RF and XGB denote Decision Tree, Random Forest and eXtreme Gradient Boosting models respectively. Included here are additional gene-specific models that were constructed but not included in the main manuscript, which focuses on the most common or highest risk CH drivers.*

| *Driver* | *n* | *Male* | *Large Clone* | *Proportion* |
|---|---|---|---|---|
| MPL | 27 | 44.44% | 51.85% | 0.01% |
| BCOR | 302 | 50.33% | 7.62% | 0.07% |
| BCORL1 | 267 | 58.43% | 7.12% | 0.06% |
| KDM6A | 211 | 42.65% | 26.07% | 0.05% |
| GNAS | 165 | 45.45% | 56.36% | 0.04% |
| CBL | 157 | 42.04% | 42.68% | 0.04% |
| SMC3 | 115 | 40.87% | 19.13% | 0.03% |
| STAG2 | 115 | 40% | 22.61% | 0.03% |
| RAD21 | 84 | 47.62% | 21.43% | 0.02% |
| EZH2 | 81 | 40.74% | 9.88% | 0.02% |
| PHF6 | 69 | 40.58% | 17.39% | 0.02% |
| RUNX1 | 54 | 53.7% | 20.37% | 0.01% |
| ZRSR2 | 50 | 66% | 44% | 0.01% |
| ETV6 | 46 | 45.65% | 28.26% | 0.01% |
| KRAS | 42 | 38.1% | 47.62% | 0.01% |

***Supplementary Table 3****: Proportions of "other" driver mutations in the UK Biobank final dataset (n=431,531).*

| *Gene* | *Number positive* | *Number in test set* | *Percentage positive* |
|---|---|---|---|
| CALR | 7 | 19 | 36.8 |
| JAK2 | 16 | 35 | 45.7 |
| SF3B1 | 5 | 51 | 9.8 |
| SRSF2 | 4 | 47 | 8.5 |
| U2AF1 | 0 | 7 | 0 |

***Supplementary Table 4****: Detection of specific driver genes by combined classifier of high-risk CH, using stringent cutoff, in the unseen test set (n = 86,306).*

**References**


1. Gu, M. *et al.* Multiparameter prediction of myeloid neoplasia risk. *Nat Genet* **55**, 1523–1530 (2023).
2. Vlasschaert, C. *et al.* A practical approach to curate clonal hematopoiesis of indeterminate potential in human genetic data sets. *Blood* **141**, 2214–2223 (2023).
3. Miller, C. A. *et al.* Failure to Detect Mutations in U2AF1 due to Changes in the GRCh38 Reference Sequence. *The Journal of Molecular Diagnostics* **24**, 219–223 (2022).
4. Danecek, P. *et al.* Twelve years of SAMtools and BCFtools. *Gigascience* **10**, giab008 (2021).
